# Coping with COVID-19 Stressors: Adverse and Protective Factors Responding to Emotions in a Chinese Sample

**DOI:** 10.1101/2021.11.30.21250895

**Authors:** Rui Xu, Xinfeng Zhang, Danni Liu, Qiang Li, Yanping Wang, Rong Jiao, Ximei Gong, Xueyan Hou, Tao Xu, Xuemei Qing, Kangxing Song, Voyko Kavcic, Shiyan Yan, Ruolei Gu, Terry Stratton, Yang Jiang

## Abstract

**Background:** The potential roles of affective responses to environmental stressors in individuals’ physical and mental health are complex and multi-faceted. This study, then, explores Chinese citizens’ emotional responses to COVID-19-related stressors and influence factors which may boost or buffer such effects.

**Methods:** From late March to early June (2020), a cross-sectional study was conducted using an anonymous online questionnaire included demographic characteristics, COVID-19-related stressors related to individuals’ daily functioning, and the self-assessed impact of protective and adverse internal factors on emotions.

**Results:** 1,662 questionnaires were received from residents in 32 Chinese provinces classified by prevalence level according to COVID-19 infections. Among the 17 positive and negative emotional responses, agglomerative hierarchical clustering revealed four subclassifications: (1) stress relations; (2) missing someone relations; (3) individual relations; and (4) social relations. Additionally, heightened regional prevalence levels positively corresponded to intensity of stress relations. Lowest intensity of social relations was found in the areas surrounding Wuhan and coastal areas. Specially, economic- and work-related stressors as well as negative self-perceptions (e.g., suppression, emotionally unstable, self-denial) implicated in negative emotions. While positive emotions were tied to demographic characteristics (e.g., high education, young age and male) and protective traits (e.g., creativity, sympathy, social responsibility), and inversely linked to relationships- and pandemic-related stressors, etc.

**Conclusion:** Associations were clearly noted among Chinese residents’ emotions to specific stressors during pandemic. Providing appropriate psychological resources/supports during future or extended public health crises may help offset the cognitive burden of individuals striving to regain an adequate level of normalcy and emotional well-being.

## Introduction

Coronavirus disease 2019 (COVID-19) have resulted in over 170 million diagnosed cases, including more than 3 million deaths worldwide (WHO, 2021). During the outbreak of COVID-19, countries imposed tight restrictions on personal behavior to halt the spread of the virus, which have also led to disruptions of routine social and economic activities, as well as health seeking behavior (Ebohon, Obienu, Irabor, Amadin, & Omoregie, 2021; Gupta & Sengupta, 2020; Shah, Quint, Nwaru, & Sheikh, 2021). The negative impact of the pandemic on world economics is already evident, as the risk of global economic crisis has been accumulating (Bank, 2021). Under these influences, the mental health of the public is of great concern during the pandemic (Alfawaz et al., 2021; Shanahan et al., 2020).

When individuals are exposed to traumatic events, their emotional responses would be modulated, which may further lead to irrational and impulsive behavior (Ceschi, Billieux, Hearn, Furst, & Van der Linden, 2014; Scott & Montgomery, 1984). Indeed, public fear-driven behaviors during the COVID-19 pandemic led to increases in doctor visits, pressures on the healthcare system, hoarding of food and daily necessities, and misuse of personal protective gear (PPG), etc. (Mahase, 2020; Oosterhoff & Palmer, 2020). Reports of irrational behaviors (e.g., delays in seeking necessary medical attention) and violence against doctors and vulnerable groups increased significantly during the restrictions (Gaballa, AlJaf, Patel, Lindsay, & Hlaing, 2020; Ghosh, 2018).

While much attention has been paid to *complex* emotional responses (e.g., anxiety and depression) to COVID-19 (Barzilay et al., 2020), *basic* emotions (e.g., happiness, sadness, fear, etc.) remain largely ignored. However, the spontaneous expression of basic emotions is more typical in “routine” social life and, subsequently, vulnerable during disruptive or anomic conditions, such as natural disaster (Y. Li et al., 2020). For instance, citizens in Croatia frequently exhibited fear, discouragement, and sadness during ten days of the COVID-19 lockdown (Dogas et al., 2020). Additionally, the public’s basic emotional expression has become a sensitive early warning indicator of infectious outbreaks such as measles, H1N1, and Ebola(Ahmed, Bath, Sbaffi, & Demartini, 2019; Du et al., 2018; Ofoghi, Mann, & Verspoor, 2016).

China has experienced the spread of COVID-19 peaked, plateaued, and declined during the first half of 2020 (Huang et al., 2020) – and infective prevalence and double-edged policies (e.g., lockdowns) subjected residents to complicated or contradictory emotional states (Jin et al., 2020; Y. Li et al., 2020). In fact, residents were forced to overcome difficulties prior to encountering social isolation and conflict (Singh & Subedi, 2020; Venkatesh, 2020; Williams, Armitage, Tampe, & Dienes, 2020). Throughout all this, the relationship of basic emotions and environmental stressors was not fully studied. Additionally, little information was available to assess perceived feelings during pandemic, although cognition was recognized to play a key role between such environmental insults and emotions. Thus, our study aimed to investigate the basic emotions and the links with self-perceived stressors, all of which facilitate governments to provide timely and effective responses to the pandemic.

## Methods

### Participants

The cross-sectional study utilized a non-random “snowball” sample designed to elicit maximum participation by consenting Chinese adults. We used an online platform of “Wenjuanxing” (https://www.wjx.cn/) and advertised by WeChat, a popular social networking platform. From late March to early June (2020), respondents to an anonymous online questionnaire came from 32 Chinese provincial districts – which were classified by prevalence level (1 = lowest, 5 = highest) according to cumulative COVID-19 infections compiled prior to the data collection deadline (see Appendix 1).

### Instrumentation

The questionnaire was devised by the authors and contained demographic information (i.e., sex, age, marital status, education) as well as self-assessed: (1) emotional responses to the COVID-19 pandemic (i.e.,9 negative emotion and 8 positive emotion); (2) environmental stressors and individuals’ daily functioning (e.g., time spent indoors); and (3) protective and adverse aspects of well-being (e.g., creative, facing death from COVID-19 infection) (see Appendix 2).

In particular, the instrument gauged respondents’ experiences with seventeen basic emotions which, based on Traditional Chinese Medicine (TCM) and recent related researches (Keltner, Sauter, Tracy, & Cowen, 2019; Ye, Cai, Cheung, & Tsang, 2019; Zhan et al., 2018). Emotions in TCM including five original types – joy (xi), anger, thinking (si), sadness, and fear – are generally considered basic, mutually-related human responses (Zhan et al., 2018). Further, each emotional type consists of varied but similar properties. For example, fear combines meanings of fearfulness and being frightened – whereas anger could be directed toward oneself or others (Praill, Gonzalez-Prendes, & Kernsmith, 2015). Additionally, thinking (si) might encompass annoyance and distracted thinking (Du, 2000). Conversely, in Chinese culture, joy (xi) represents positive emotions such as happiness/joy, comfort/relaxation, a sense of accomplishment, and safety (Dictionary Editing Room, 2011). All emotions could result in response to a pandemic, which were assessed on a 4-point Likert-type scale ranging from 0 (“Not at all”) to 3 (“A great deal”).

Environmental stressors assessed during the COVID-19 pandemic focused on disruptions to daily life, difficulties, or personal experiences resulting from infection or infective prevalence. Subjects also assessed protective and adverse internal factors impacting their emotions. All items were derived from established Chinese COVID-19-related guidelines for psychological assistance/service/counseling provided by various resources and organizations (Ma, 2020; Shen & Wang, 2020; Wu & Zhang, 2020; Zhao & Liu, 2020).

### Statistical Analysis

#### Agglomerative hierarchical clustering

As a data reduction technique, and to identify underlying data structures, we used the maximum coefficient method of agglomerative hierarchical clustering (James & Tibshirani, 2013). In this iterative process, items initially treated as individual clusters are progressively collapsed based on Pearson correlation coefficients (Casella & Berger, 2001). The correlation matrix is then recalculated and the maximum coefficient again identified between the new and uncombined clusters. This is repeated until all items are grouped together into a single cluster or the procedure exceeds the specified number of iterations.

#### LASSO Regression

A major challenge to classifying multiple objects within a singular construct is a large intra-class variation that typically exists. The LASSO approach to parameter estimation alters the model-fitting process to select only a subset of covariates which produce an optimally predictive and interpretable model (Tibshirani, 2011).This goal is achieved by utilizing a regularization process which, when applied across all predictors in a multivariate model, allows the contributions of some coefficients to be reduced to zero and, hence, completely removed from the model. This advantage, among others, distinguishes LASSO from similar techniques (e.g., ridge regression) designed to avoid model over-fitting by reducing residual “noise” (http://wavedatalab.github.io/machinelearningwithr/post4.html).

As a parallel to designating measures as dependent or independent variables, LASSO uses machine learning (ML) vernacular to reference inputs (“features”) and outputs (“labels”). More specifically, “features” are properties of the data used to “train” or develop the model-fitting algorithm, while “labels” are the resulting output returned after computing.

Based on this framework, our training sample included 65 features in the four-part feature set (including environment stressors, protective and adverse internal factors, and demographic information, see Appendix 1), while the label set was defined a certain emotion cluster. Normalizing the feature variables to obtain a standard sample set, each standard sample had a feature vector (X) and a label variable (Y) upon which the LASSO regression model was built. The number of features ultimately included in the final model is selected by a cross-validation method and the parameters obtained. We chose 20% of the overall data as the test set used to gauge performance of the selected model.

A linear regression that models the response variable y using a set of *d* features, ***x***_***i***_ ***=*** [1, *x*_*i*1,_ …, *x*_*id*_]^*T*^,for *i*th case can be expressed as:

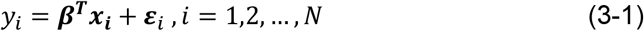

where ***β =*** [*β*_0_, *β*_1,_ …, *β*_*d*_]^*T*^ is a vector, each of its components represents a coefficient of the linear model, and ***ε***_*i*_ is the residual term for the *i*th case representing remaining unexplained variance. For a set of N cases, the model can be rewritten in the form of the following matrix:

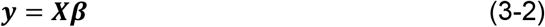

where ***y =*** [*y*_1,_ …, *y*_*N*_]^*T*^, and ***X*** is an *N* × *d* matrix with features in columns, as follows:

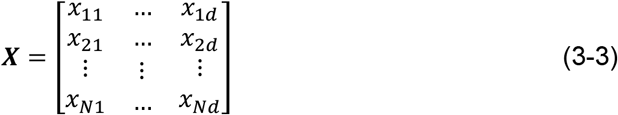

where entry *x*_*ij*_ is the *j*th feature for the *i*th case.

Ordinary least squares (OLS) is often used to generate linear model coefficients *β* as:

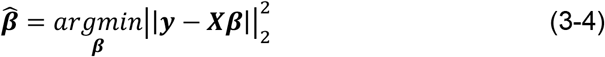

With multiple potential predictors, a smaller, more parsimonious subset of “features” is sought which exhibits the strongest effects. Toward this end, OLS estimates are suboptimal to regularization approaches which impose “penalties” on certain coefficients to avoid model overfitting. As mentioned above, LASSO regression shrinks some coefficients and sets others to 0 – obtaining the optimal solution of the following equation via a penalty term:

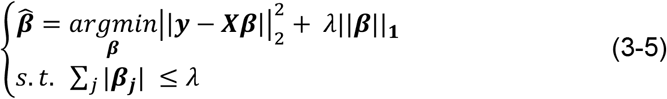

where 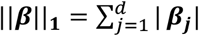, the first part represents the goodness of the model fitting, and the second part represents the parameter penalty. Here, λ ≥ 0 is a complexity parameter that controls the amount of shrinkage as coefficients are reduced or set to zero: the larger the value of λ, the greater the amount of shrinkage. The smaller the regularization parameter *λ*, the less punitive the model and the more features it retains. Conversely, increases in *λ* results in reduced numbers of features.

Additionally, Kruskal-Wallis ANOVA were adopted to explore the differences of each emotional cluster (i.e., positive, negative; stress relations, missing sb. relations, individual relations, social relations) among five regional prevalence levels of COVID-19 infective (1 = lowest, 5 = highest). Post hoc comparisons were also conducted.

A critical p value of < .05 was set for all inferential tests, and analyses were conducted using Python (https://www.python.org/) and R software platforms (https://www.python.org/).

## Results

### Sample characteristics

Nineteen (19) of 1,681 the questionnaires were excluded due to incomplete responses or participants not living in China – yielding a final, analyzable sample of 1,662. The “typical” respondent was female (60.5%, n = 1,006) and 30.7 years of age (SD = 10.3). Married (49.0%, n = 815) and single (49.2%, n = 817) respondents, respectively, each accounted for virtually one-half of the sample. Most respondents (63.2%) had bachelor’s degrees or above, and the prevalence rate of COVID-19 infection across the five regions ranged from ∼13.4 to 28.0%. Sample demographics are summarized in Table 1 – along with reported emotional reactions and influential factors (e.g., situational stressors, protective/adverse internal factors). Descriptive statistics of the latter are presented below.

**Table 1.** Descriptive statistics of study variables (n=1,662)

| **Variables** | **Frequency (%)** | **Mean (SD)** |
| --- | --- | --- |
| **Demographic information** |  |  |
| Age (years) |  | 30.66 (10.3) |
| Sex |  |  |
| Male | 656 (39.5%) |  |
| Female | 1006 (60.5%) |  |
| Marital status |  |  |
| Single | 817 (49.2%) |  |
| Married | 815 (49.0%) |  |
| Divorced | 25 (1.6%) |  |
| Widowed | 5 (0.3%) |  |
| Education |  |  |
| Senior high school/technical secondary school and below | 171 (10.3%) |  |
| Associate degree in college | 440 (26.5%) |  |
| Bachelor's degree | 835 (50.2%) |  |
| Master's degree or greater | 216 (13.0%) |  |
| Respondents by Region |  |  |
| 5 (Highest Prevalence) (1 province) |  | 291(17.5) |
| 4 (5 provinces) |  | 223(13.4) |
| 3 (9 provinces) |  | 465 (28.0) |
| 2 (10 provinces) |  | 344(20.7) |
| 1 (Lowest Prevalence)) (7 provinces) |  | 339 (20.4) |
| **Emotions** |  |  |
| For two classification |  |  |
| Negative emotions (V1 ~ V9) |  | 1.91 (0.6) |
| Positive emotions (V10 ~ V17) |  | 2.33 (0.6) |
| For four classification |  |  |
| Stress-related NE (V1 ~ V8) |  | 1.87 (0.7) |
| Missing (V9) |  | 2.18 (0.7) |
| Individual-related PE (V10 ~ V14) |  | 2.57 (0.8) |
| Social-related PE (V15 ~ V17) |  | 2.20 (1.0) |
| **Influence factors** |  |  |
| Situational stressors |  | 5.20 (4.0) |
| Protective internal factors |  | 5.27 (3.3) |
| Adverse internal factors |  | 3.54 (3.5) |

Note: SD, Standard Deviation.

### Emotional Clusters

Seventeen (17) emotional responses to COVID-19 were divided into two categories reflecting positive and negative emotions and, subsequently, four nested subclassifications: For negative emotions, the emotion “missing someone who is close to you” was distanced from those related to *stress* (e.g., sadness, fear/fright, anger toward oneself or others, worry, annoyance and distracted thinking due to anger). Positive emotions, in contrast, were classified as individual- (e.g., happiness, relaxation, comfort, safety and accomplishment) or social-relations (e.g., emotionally inspired, empathy, moved by sb. or sth.). The associated correlation matrix and cluster dendrogram are shown below in Figures 1A and 1B, respectively.

**Figure 1.**
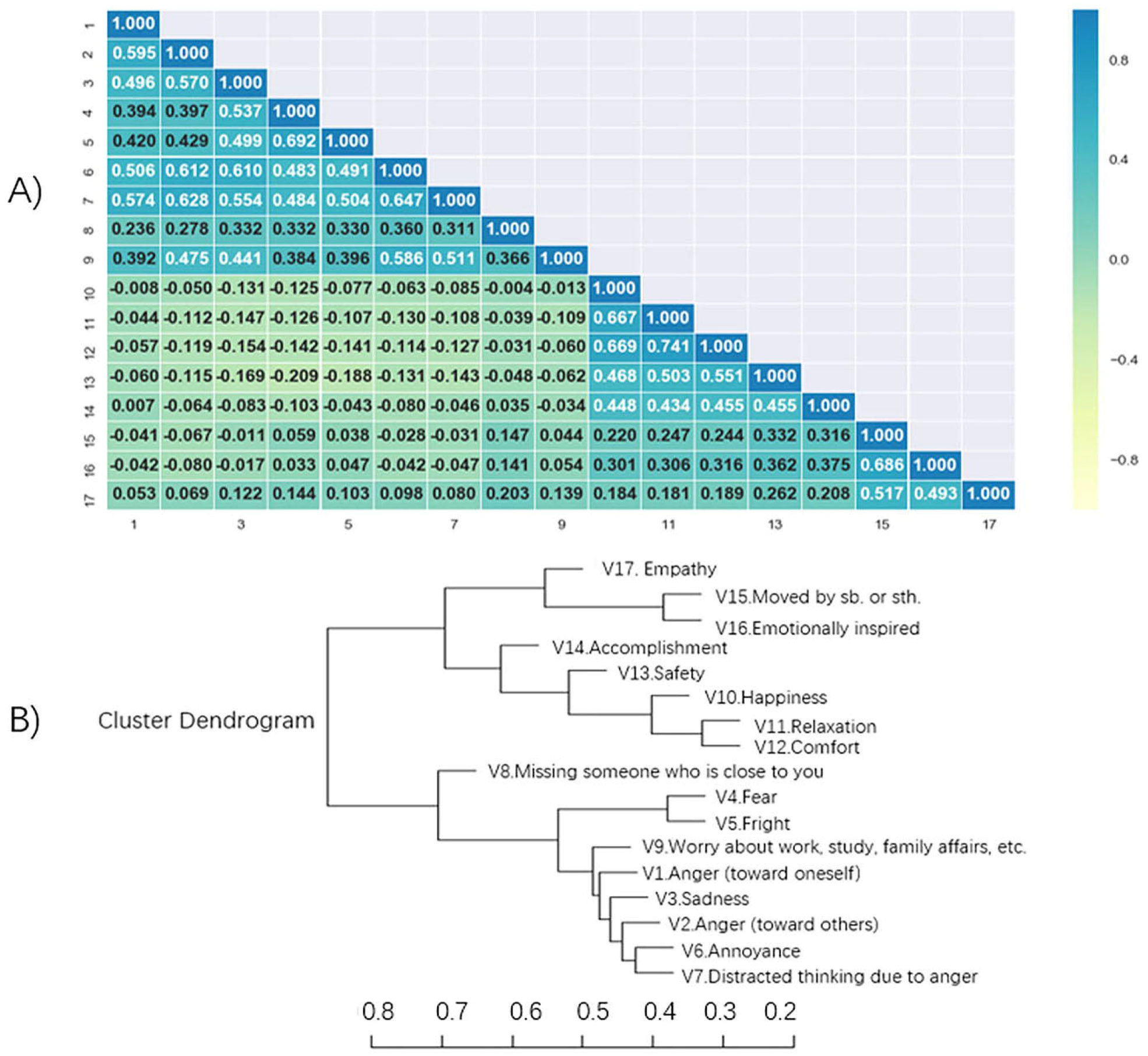
The Emotional Correlation Matrix and Cluster Dendrogram. A) Emotional correlation matrix was calculated by Pearson correlation coefficients among the seventeen emotions. B) Agglomerative hierarchical clustering was adopted the maximum coefficient method.

### LASSO Feature Selection: Emotions

LASSO regression was adopted to select predictors for emotional clusters. Based on performance parameters, the models of feature selection were acceptable for emotions classified as negative (***λ***_**0**_= 0.0228, r2 = 0.5524, MSE (Mean Squared Error) = 0.1910, numbers of predictors = 34) and positive (***λ***_**0**_= 0.0173, r2 = 0.1712, MSE =0.2841, numbers of predictors = 32).

A cross-validation process intended to find the optimal ***λ*** value (***λ***_**0**_) - designating the most parsimonious model with a minimum average MSE (see Figure 2A). As ***λ*** increases, more coefficients are set to zero - leading to a sparser model (see Figure 2B).

**Figure 2.**
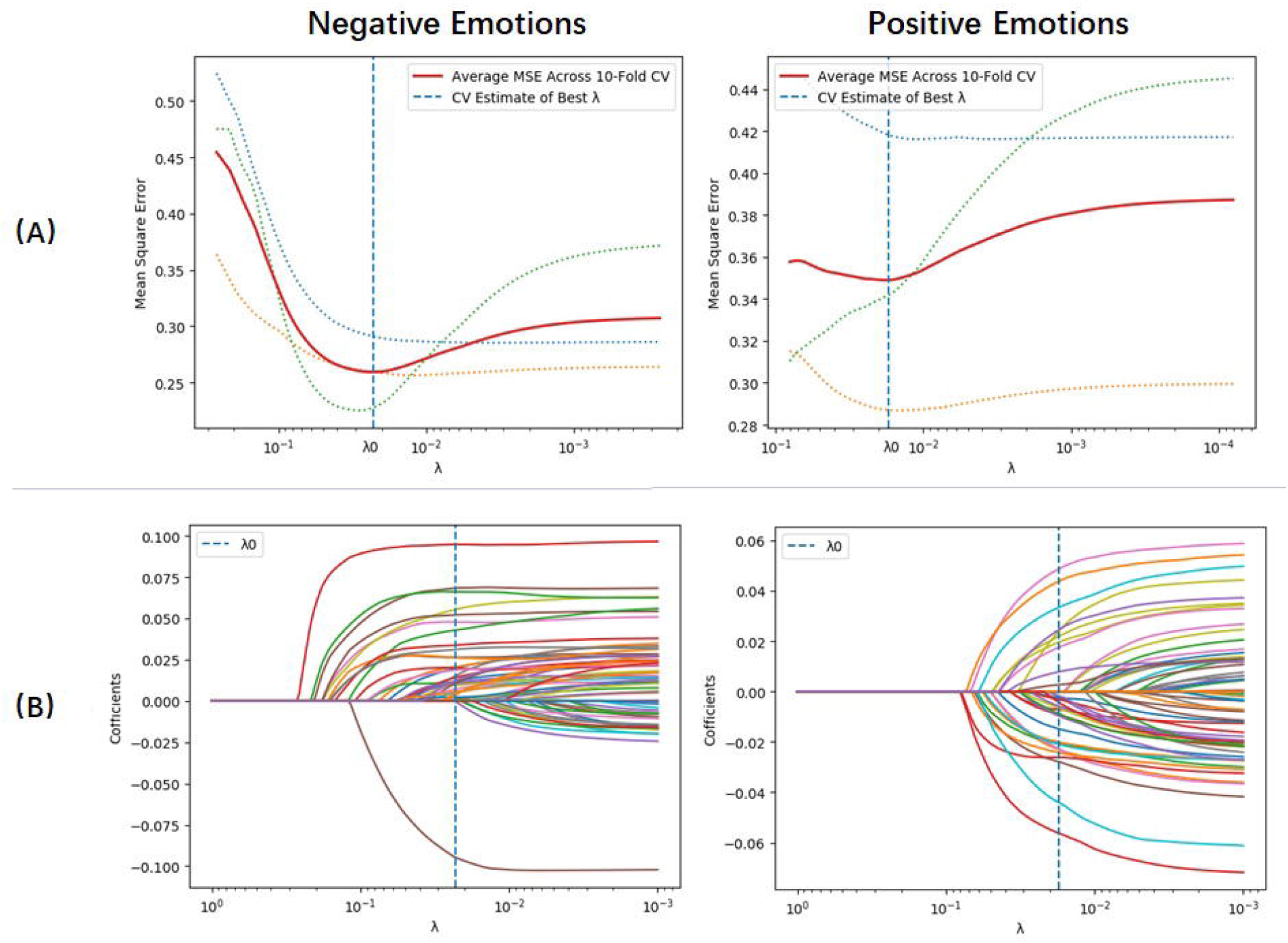
The Performance Parameters of LASSO Feature Selection. (A) Average MSE of the LASSO models as a function of the regularization parameter ***λ***. The optimal penalty, determined by 3-fold cross-validation, is the value of ***λ*** that minimizes the MSE (shown as a vertical dashed line). (B) Trace plot showing non-zero model coefficients as a function of the regularization parameter ***λ***. As ***λ*** increases to the left, coefficients are set to zero and removed from the model.

Corresponding coefficients for the emotional models with all relevant predictors are reported in Figure 3. Results showed that the predictors of negative emotions included a tendency to suppress emotions, emotional instability, self-denial, vulnerability to impact, economic problems, fatigue/sleepiness, work-related problems, unequal treatment, feeling limited/constrained, lacking confidence etc. In contrast, positive emotions were driven by creativity, sympathy, social responsibility, high education etc., while factors, such as tendency to suppress emotions, age, intimate partner relationship, social isolation and facing death from COVID-19, had the opposite effect.

**Figure 3.**
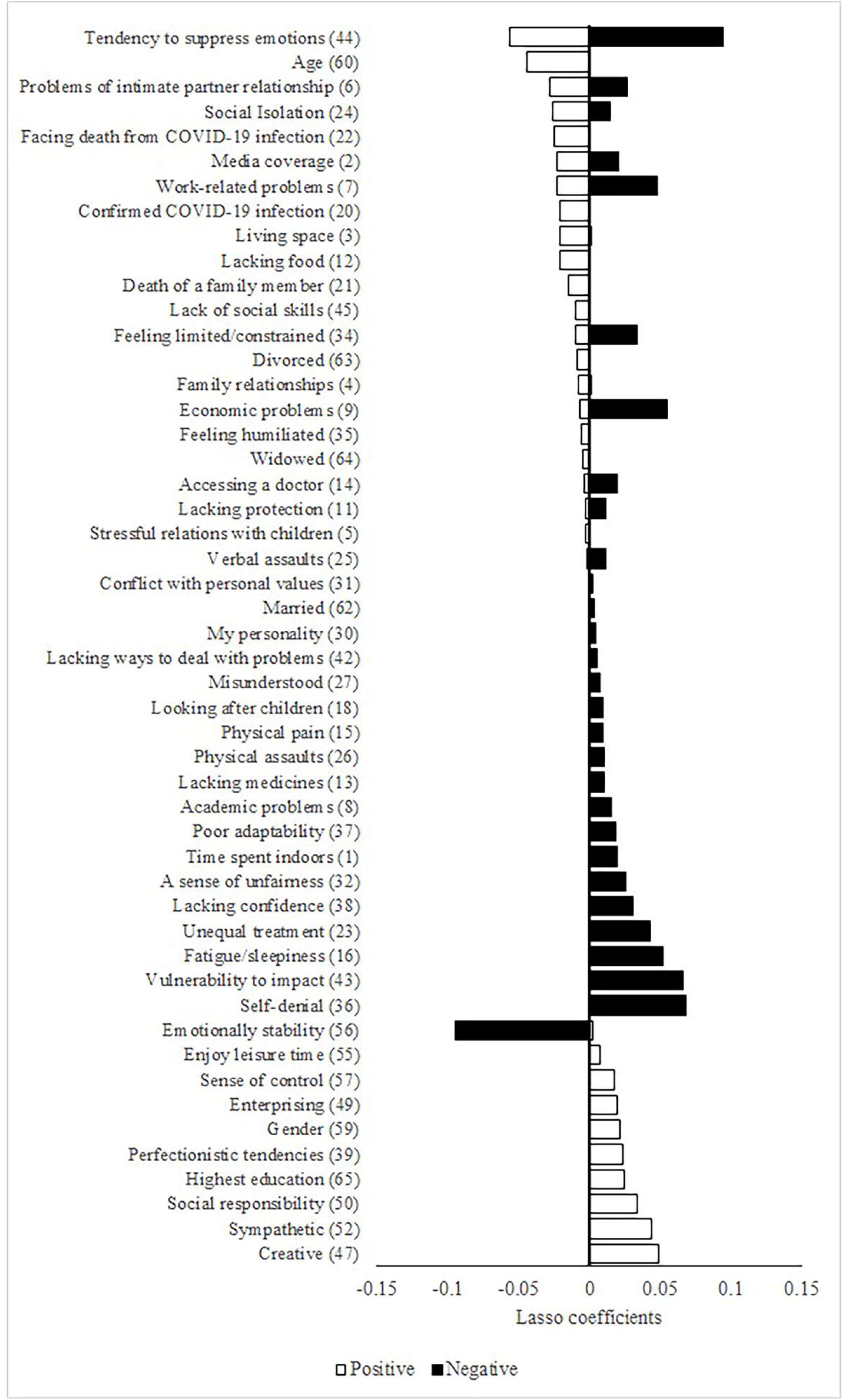
Beta coefficients for predictors of LASSO Feature Selection for Negative and Positive Emotions. X-axis exhibited the coefficients of LASSO regression. Y-axis showed stressors whose coefficients weren’t zero.

### Emotional Response by Regional Level of Infection

Significant differences in emotional clusters among residents from regions with varying levels of COVID-19 infective prevalence were found (see Table 2), with pairwise, post hoc comparisons (see Table 3) showing positive emotions in the highest prevalence region were lower than the medium or lowest prevalence regions. Significant differences in negative emotions were found among all pairs of regions except higher vs. medium. For nested subclassifications, individual relations of positive emotions tended to be lowest among respondents located in the highest prevalence region. Social relations found to be lowest in the higher prevalence region, in where were significantly lower than medium and lowest regions. Regarding negative emotions, stress relations varied significantly among respondents from the various regions – with the exception of lower vs. higher and higher vs. medium. Lastly, those from lower prevalence regions reported significantly lower emotional responses related to missing sb. relations.

**Table 2.** Emotional Differences Among Regions with Levels of COVID-19 Infective Prevalence. (Mean ± SE, 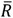)

| Risk Regions | For two classification |  | For four classification |  |  |  |
| --- | --- | --- | --- | --- | --- | --- |
|  | Negative Emotions | Positive Emotions | Stress Relations | Missing sb. Relations | Individual Relations | Social Relations |
| 5 (Highest) | 2.26±0.03 (1108.50) | 2.22±0.03 (744.37) | 2.24±0.03 (1190.46) | 2.47±0.06 (955.57) | 2.01±0.03 (710.22) | 2.56±0.04 (819.86) |
| 4 | 1.94±0.04 (865.69) | 2.29±0.04 (805.33) | 1.89±0.04 (853.30) | 2.37±0.07 (909.15) | 2.22±0.04 (868.00) | 2.40±0.05 (731.33) |
| 3 | 1.97±0.03 (869.16) | 2.34±0.03 (855.96) | 1.93±0.03 (867.25) | 2.26±0.04 (859.82) | 2.20±0.03 (843.69) | 2.58±0.04 (840.33) |
| 2 | 1.78±0.03 (730.64) | 2.32±0.03 (816.75) | 1.75±0.03 (739.09) | 2.03±0.05 (745.23) | 2.19±0.04 (839.93) | 2.54±0.05 (820.80) |
| 1 (Lowest) | 1.65±0.03 (621.91) | 2.43±0.03 (904.92) | 1.61±0.03 (623.29) | 1.96±0.05 (722.62) | 2.26±0.04 (886.32) | 2.71±0.04 (906.13) |
| Kruskal-Wallis H | 181.35 | 19.83 | 178.00 | 60.44 | 25.06 | 18.76 |
| *Adj. Sig.* | < 0.001 | 0.001 | < 0.001 | < 0.001 | < 0.001 | 0.001 |

Note: SD, Standard Error; $\bar{R}$ , Average Rank.

**Table 3.** Post Hoc for Emotional Differences Among Regions with Infective Prevalence Levels.

|  | **Negative**<br>**Emotions** | **Positive**<br>**Emotions** | **Stress**<br>**Relations** | **Missing sb.**<br>**Relations** | **Individual**<br>**Relations** | **Social**<br>**Relations** |
| --- | --- | --- | --- | --- | --- | --- |
| **Sample1-Sample2** | *Adj_p* | *Adj_p* | *Adj_p* | *Adj_p* | *Adj_p* | *Adj_p* |
| 1-2 | 0.030 | 0.161 | 0.016 | 1.000 | 1.000 | 0.190 |
| 1-3 | 0.000 | 1.000 | 0.000 | 0.000 | 1.000 | 0.527 |
| 1-4 | 0.000 | 0.158 | 0.000 | 0.000 | 1.000 | 0.000 |
| 1-5 | 0.000 | 0.000 | 0.000 | 0.000 | 0.000 | 0.232 |
| 2-3 | 0.000 | 1.000 | 0.002 | 0.005 | 1.000 | 1.000 |
| 2-4 | 0.000 | 1.000 | 0.055 | 0.000 | 1.000 | 0.286 |
| 2-5 | 0.000 | 0.576 | 0.000 | 0.000 | 0.006 | 1.000 |
| 3-4 | 1.000 | 1.000 | 1.000 | 1.000 | 1.000 | 0.049 |
| 3-5 | 0.000 | 0.018 | 0.000 | 0.053 | 0.002 | 1.000 |
| 4-5 | 0.000 | 1.000 | 0.000 | 1.000 | 0.002 | 0.364 |

Note: Prevalence level according to numbers of COVID-19 infections (1 = lowest, 5 = highest)

## Discussion

Our study investigated public emotions and related influence factors associated with the COVID-19 pandemic in China. Negative emotions were consistently confirmed during this time (see also (Du et al., 2018), with previous studies revealing variation in emotional strength and category across phases of the pandemic (CITE). For instance, an analysis of Twitter posts during a recent Ebola outbreak found different basic emotional responses prior to and after key incidents (Ofoghi et al., 2016). A recent study also showed that Chinese residents’ emotional expression via Weibo (a Chinese version of Twitter) was strongest before the actual peak of COVID-19 outbreak and declined thereafter (Y. Li et al., 2020). Additionally, our study revealed strength of negative emotional response was sensitive to gradient regional prevalence levels by investigated data. Consistently, significant correlations were found between regional cumulative COVID-19 cases and Web searches by Italian netizens including the generic terms “fear” and “anxiety” (Rovetta & Castaldo, 2020). However, missing sb. relations didn’t have such sensitivity, showing significantly lower intensity in lower prevalence regions than other prevalence regions. Additionally, lowest social relations showed in the higher prevalence region (areas surrounding Wuhan and coastal areas), which may be due to the typhoon eye’ effect. In line with recent works, working adults’ distance to the epicenter of Wuhan had an inverted U-shaped relationship with their burnout (Zhang, Huang, & Wei, 2020).

The differences of situational stressors and self-perception associated with emotional reactions were further analyzed. Specifically, negative emotions were triggered largely by the economic and work-related problems and negative self-images, while positive emotions connected with relationship- and epidemic-related stressors, personal characteristics and demographic variables). Indeed, besides the prevalence of viral infection, COVID-19 exacted a global economic toll and social problems of unprecedented scope, magnitude, and duration (Nicola et al., 2020; Ramos Perkis et al., 2020). Strict control measures have been instituted and changed public normal routines with shutting down of businesses, industries, and schools. Uncertainty about opportunities of employment and academic has been increased worldwide, even though pandemic has been controlled in China (Wilson et al., 2020). Thus, public emotions fluctuated largely as a consequence of unfulfilled needs of survival- and self-development, when facing dilemma of income decline, unemployment and business closes.

Self-perception played a key role for emotions revealed in our study. The frequent tendency to suppress certain affective responses all emotions, which showed the more emotions are suppressed, the easier it is to cause negative emotions and harder to form positive emotions in our study. Literatures illustrated emotional expression potentially impacted mental well-being, psychophysiological responses and interaction behaviors (Nagulendran, Norton, & Jobson, 2020; Waters, Karnilowicz, West, & Mendes, 2020). A recent study showed that critical work, health and family engagement was worse with increased suppression of fear and apprehension about COVID-19 infection (Trougakos, Chawla, & McCarthy, 2020). Additionally, self-perception, as a core process of coping with stress, had a practical utility in explaining the public’s emotion and behavior (J. B. Li et al., 2020). For instance, negative self-images (e.g., self-denial, emotionally unstable, lacking confidence) frequently manifested by mood disorders, hindering the function of emotional regulation, resulted in and deepen the degree of negative emotions (Kawashima et al., 2016; Laghi, Bianchi, Pompili, Lonigro, & Baiocco, 2018; Masuda et al., 2017). Such adverse internal factors (i.e., easy to be affected, feeling constrained) as well as stressors (media coverage and a long-time spent indoors) together contributed to negative affection, which was consistently found in recent works (Basch et al., 2020).

The oldest conjectures was confirmed that happiness depends not just on absolute things but inherently on comparisons with other people (Clark & Oswald, 2002). Evidence of comparison effects was also revealed in our results, that was high intensity of positive emotions related with experiencing fewer and not serious negative incidents, such as intimate partner relationship, social isolation, and facing death from COVID-19 infection. In line with previous studies, issues of health and interpersonal relationships were especially concerned by residents, when encountering with disaster situations (Pieh, T, Budimir, & Probst, 2020). For instance, a survey found survivors’ mental and physical health and neighborhood connectedness were significantly correlated with happiness in the fifth years after the Tohoku Earthquake Tsunami in Japan (Sun & Yan, 2019). Social isolation significantly predicted poor mental health associated with COVID-19 policy, particularly for elders, or individuals in low-paid or precarious employment (Kim & Jung, 2020; Williams et al., 2020).

Positive emotional state and self-perceptions played protective roles for residents in encounter of difficulties. As previous studies confirmed, positive emotions serving as the ultimate target or a moderator, helped people to cope with negative events (Waugh, 2020). Similarity, positive self-perceptions associated with high self-esteem could minimize influence of negative information (Showers, 1992). It was worth to notice that creativity made a great contribution to positive emotional response to the COVID-19. To tackle the unanticipated difficulties and intricate problems during pandemic, interaction between creativity and positive feelings may play an important role to promote task completion (Bang & Reio, 2017), as well as to change attitudes and expected directions (Walsh, Chen, Hacker, & Broschard, 2008). Additionally, protective factors also highlighted some self-assessed characteristics which were related with social members (e.g., sympathy and socially responsible). Consistently, conscientiousness and openness were found as a protective factor for sadness, depression, stress and tension caused by stress (Schlee et al., 2020). Higher sympathy associated with higher prosocial behavior, which was also found to be the positive link between sadness regulation and prosocial behavior and to mediate by higher sympathy and trust (Song, Colasante, & Malti, 2018). Lastly, high education, young age and male contributed to positive emotional state, which was proved to potentially mitigate and regulate the psycho-emotional toll (Banati, Jones, & Youssef, 2020; Greiner, Muller, Norris, Ng, & Sangha, 2019).

The limitation of the study is that most responders in our sample haven’t experience traumatic events, such as confirmed COVID-19 infection. Thus, they reported lots of maladjustment for changes of routine life. Those who and whose family members have fallen illness due to virus infections, may show more intensity of negative emotions and different stressors and influence factors. In addition, we did not ask participants to report their quality of life, thus the potential influence of emotion on subjective well-being has not been fully understood.

## Conclusion

Negative emotional response was sensitive to regional prevalence levels of COVID-19. Economic and work-related problems largely contributed to negative emotions and relationship- and epidemic-related stressors largely affected positive emotions. Negative self-images (i.e., emotional expression, self-denial, emotionally unstable, lacking confidence) were adverse factors for emotions. Protective factors for emotions included creativity, social-orient traits, as well as high education, young age and male. Future study should investigate emotional supports/measures based on interactions among particular stressors and cognitive burden, as well as protective factors.

## Data Availability

The data and material can be accessed from

## Authors’ contribution

RX, TS, and YJ designed the study. RX, TS, SY and YJ wrote the study protocol. QL, YW, RJ, XG, XH, TX, JY, and KS collected the data. XZ, DL and RX carried out the analysis with support from SY, TS, VK and YJ. RX and TS wrote the draft of the manuscript. TS, DL, SY, XZ, RG, VK and YJ revised the draft. All authors contributed to the final version of the manuscript.

## Acknowledgements

Not applicable

## Reference

Ahmed, W., Bath, P. A., Sbaffi, L., & Demartini, G. (2019). Novel insights into views towards H1N1 during the 2009 Pandemic: a thematic analysis of Twitter data. Health Info Libr J, 36(1), 60–72. doi:10.1111/hir.12247

Alfawaz, H. A., Wani, K., Aljumah, A. A., Aldisi, D., Ansari, M. G. A., Yakout, S. M., … Al-Daghri, N. M. (2021). Psychological well-being during COVID-19 lockdown: Insights from a Saudi State University’s Academic Community. J King Saud Univ Sci, 33(1), 101262. doi: 10.1016/j.jksus.2020.101262

Banati, P., Jones, N., & Youssef, S. (2020). Intersecting Vulnerabilities: The Impacts of COVID-19 on the Psycho-emotional Lives of Young People in Low- and Middle-Income Countries. Eur J Dev Res, 1–26. doi:10.1057/s41287-020-00325-5

Bang, H., & Reio, T. G., Jr. (2017). Personal Accomplishment, Mentoring, and Creative Self-Efficacy as Predictors of Creative Work Involvement: The Moderating Role of Positive and Negative Affect. J Psychol, 151(2), 148–170. doi:10.1080/00223980.2016.1248808

Barzilay, R., Moore, T. M., Greenberg, D. M., DiDomenico, G. E., Brown, L. A., White, L. K., … Gur, R. E. (2020). Resilience, COVID-19-related stress, anxiety and depression during the pandemic in a large population enriched for healthcare providers. Transl Psychiatry, 10(1), 291. doi:10.1038/s41398-020-00982-4

Basch, C. H., Hillyer, G. C., Erwin, Z. M., Mohlman, J., Cosgrove, A., & Quinones, N. (2020). News coverage of the COVID-19 pandemic: Missed opportunities to promote health sustaining behaviors. Infect Dis Health, 25(3), 205–209. doi:10.1016/j.idh.2020.05.001

Casella, G., & Berger, R. L. (2001). Statistical Inference. Duxbury Press, 169–172.

Ceschi, G., Billieux, J., Hearn, M., Furst, G., & Van der Linden, M. (2014). Trauma exposure interacts with impulsivity in predicting emotion regulation and depressive mood. Eur J Psychotraumatol, 5. doi:10.3402/ejpt.v5.24104

Clark, A. E., & Oswald, A. J. (2002). A simple statistical method for measuring how life events affect happiness. Int J Epidemiol, 31(6), 1139-1144; discussion 1144-1146. doi:10.1093/ije/31.6.1139

Dictionary Editing Room, Institute of Linguistics, Chinese Academy of Social Sciences. (2011). Xinhua dictionary (11th edition). Beijing: Commercial Press, 532.

Dogas, Z., Lusic Kalcina, L., Pavlinac Dodig, I., Demirovic, S., Madirazza, K., Valic, M., & Pecotic, R. (2020). The effect of COVID-19 lockdown on lifestyle and mood in Croatian general population: a cross-sectional study. Croat Med J, 61(4), 309–318.

Du, J., Tang, L., Xiang, Y., Zhi, D., Xu, J., Song, H. Y., & Tao, C. (2018). Public Perception Analysis of Tweets During the 2015 Measles Outbreak: Comparative Study Using Convolutional Neural Network Models. J Med Internet Res, 20(7), e236. doi:10.2196/jmir.9413

Du, W. (2000). [The essence and clinical implication of “thinking”]. Journal of Nanjing University of TCM (social sciences), 1(2), 100–102.

Ebohon, O., Obienu, A. C., Irabor, F., Amadin, F. I., & Omoregie, E. S. (2021). Evaluating the impact of COVID-19 pandemic lockdown on education in Nigeria: Insights from teachers and students on virtual/online learning. Bull Natl Res Cent, 45(1), \p76. doi: 10.1186/s42269-021-00538-6

Gaballa, S., AlJaf, A., Patel, K., Lindsay, J., & Hlaing, K. M. (2020). COVID-19 Fears May Be Worse Than the Virus: A Case of Cardiogenic Shock Secondary to Post-Myocardial Infarction Ventricular Septum Rupture. Cureus, 12(6), e8809. doi:10.7759/cureus.8809

Ghosh, K. (2018). Violence against doctors: A pandemic in the making. Eur J Intern Med, 50, e9–e10. doi:10.1016/j.ejim.2017.08.017

Greiner, E. M., Muller, I., Norris, M. R., Ng, K. H., & Sangha, S. (2019). Sex differences in fear regulation and reward-seeking behaviors in a fear-safety-reward discrimination task. Behav Brain Res, 368, 111903. doi:10.1016/j.bbr.2019.111903

Gupta, P., & Sengupta, A. (2020). A qualitative analysis of social scientists’ opinions on socioeconomic and demographic implications of the lockdown during COVID-19 in India. J Public Aff, e2531. doi: 10.1002/pa.2531

Huang, C., Wang, Y., Li, X., Ren, L., Zhao, J., Hu, Y., … Cao, B. (2020). Clinical features of patients infected with 2019 novel coronavirus in Wuhan, China. Lancet, 395(10223), 497–506. doi:10.1016/S0140-6736(20)30183-5

James, G., & Tibshirani, R. (2013). An Introduction to Statistical Learning with Applications in R. Springer, 393,399.

Jin, Z., Zhao, K. B., Xia, Y. Y., Chen, R. J., Yu, H., Tamunang Tamutana, T., … Park, G. Y. (2020). Relationship Between Psychological Responses and the Appraisal of Risk Communication During the Early Phase of the COVID-19 Pandemic: A Two-Wave Study of Community Residents in China. Front Public Health, 8, 550220. doi:10.3389/fpubh.2020.550220

Kawashima, C., Tanaka, Y., Inoue, A., Nakanishi, M., Okamoto, K., Maruyama, Y., … Akiyoshi, J. (2016). Hyperfunction of left lateral prefrontal cortex and automatic thoughts in social anxiety disorder: A near-infrared spectroscopy study. J Affect Disord, 206, 256–260. doi:10.1016/j.jad.2016.07.028

Keltner, D., Sauter, D., Tracy, J., & Cowen, A. (2019). Emotional Expression: Advances in Basic Emotion Theory. J Nonverbal Behav, 43(2), 133–160. doi:10.1007/s10919-019-00293-3

Kim, H. H., & Jung, J. H. (2020). Social Isolation and Psychological Distress During the COVID-19 Pandemic: A Cross-National Analysis. Gerontologist. doi:10.1093/geront/gnaa168

Laghi, F., Bianchi, D., Pompili, S., Lonigro, A., & Baiocco, R. (2018). Metacognition, emotional functioning and binge eating in adolescence: the moderation role of need to control thoughts. Eat Weight Disord, 23(6), 861–869. doi:10.1007/s40519-018-0603-1

Li, J. B., Yang, A., Dou, K., Wang, L. X., Zhang, M. C., & Lin, X. Q. (2020). Chinese public’s knowledge, perceived severity, and perceived controllability of COVID-19 and their associations with emotional and behavioural reactions, social participation, and precautionary behaviour: a national survey. BMC Public Health, 20(1), 1589. doi:10.1186/s12889-020-09695-1

Li, Y., Zeng, Y., Liu, G., Lu, D., Yang, H., Ying, Z., … Song, H. (2020). Public awareness, emotional reactions and human mobility in response to the COVID-19 outbreak in China - a population-based ecological study. Psychol Med, 1–8. doi:10.1017/S003329172000375X

Mahase, E. (2020). Covid-19: hoarding and misuse of protective gear is jeopardising the response, WHO warns. BMJ, 368, m869. doi:10.1136/bmj.m869

Masuda, K., Nakanishi, M., Okamoto, K., Kawashima, C., Oshita, H., Inoue, A., … Akiyoshi, J. (2017). Different functioning of prefrontal cortex predicts treatment response after a selective serotonin reuptake inhibitor treatment in patients with major depression. J Affect Disord, 214, 44–52. doi:10.1016/j.jad.2017.02.034

Ma, X. (2020). Guidelines to Public Psychological Self-help and Counselling for COVID-19. Beijing, CHN: People’s Medical Publishing House.

Nagulendran, A., Norton, P. J., & Jobson, L. (2020). Investigating cultural differences in the effects of expressive suppression when processing traumatic distressing material. Behav Cogn Psychother, 48(6), 658–671. doi:10.1017/S1352465820000508

Nicola, M., Alsafi, Z., Sohrabi, C., Kerwan, A., Al-Jabir, A., Iosifidis, C., … Agha, R. (2020). The socio-economic implications of the coronavirus pandemic (COVID-19): A review. Int J Surg, 78, 185–193. doi:10.1016/j.ijsu.2020.04.018

Ofoghi, B., Mann, M., & Verspoor, K. (2016). Towards Early Discovery of Salient Health Threats: A Social Media Emotion Classification Technique. Pac Symp Biocomput, 21, 504–515.

Oosterhoff, B., & Palmer, C. A. (2020). Attitudes and Psychological Factors Associated With News Monitoring, Social Distancing, Disinfecting, and Hoarding Behaviors Among US Adolescents During the Coronavirus Disease 2019 Pandemic. JAMA Pediatr. doi:10.1001/jamapediatrics.2020.1876

Pieh, C. TO. R., Budimir, S., & Probst, T. (2020). Relationship quality and mental health during COVID-19 lockdown. PLoS One, 15(9), e0238906. doi:10.1371/journal.pone.0238906

Praill, N., Gonzalez-Prendes, A. A., & Kernsmith, P. (2015). An Exploration of the Relationships between Attitudes Towards Anger Expression and Personal Style of Anger Expression in Women in the USA and Canada. Issues Ment Health Nurs, 36(6), 397–406. doi:10.3109/01612840.2014.992082

Ramos Perkis, J. P., Achurra Tirado, P., Raykar, N., Zinco Acosta, A., Munoz Alarcon, C., Puyana, J. C., & Ottolino Lavarte, P. (2020). Different Crises, Different Patterns of Trauma. The Impact of a Social Crisis and the COVID-19 Health Pandemic on a High Violence Area. World J Surg. doi:10.1007/s00268-020-05860-0

Rovetta, A., & Castaldo, L. (2020). The Impact of COVID-19 on Italian Web Users: A Quantitative Analysis of Regional Hygiene Interest and Emotional Response. Cureus, 12(9), e10719. doi:10.7759/cureus.10719

Schlee, W., Holleland, S., Bulla, J., Simoes, J., Neff, P., Schoisswohl, S., … Langguth, B. (2020). The Effect of Environmental Stressors on Tinnitus: A Prospective Longitudinal Study on the Impact of the COVID-19 Pandemic. J Clin Med, 9(9). doi:10.3390/jcm9092756

Scott, V. L., & Montgomery, P. H. (1984). Urban violence: pandemic of trauma. Emerg Med Serv, 13(5), 18, 20-11.

Shah, S. A., Quint, J. K., Nwaru, B. I., & Sheikh, A. (2021). Impact of COVID-19 national lockdown on asthma exacerbations: interrupted time-series analysis of English primary care data. Thorax. doi: 10.1136/thoraxjnl-2020-216512

Shanahan, L., Steinhoff, A., Bechtiger, L., Murray, A. L., Nivette, A., Hepp, U., … Eisner, M. (2020). Emotional distress in young adults during the COVID-19 pandemic: evidence of risk and resilience from a longitudinal cohort study. Psychol Med, 1–10. doi: 10.1017/S003329172000241X

Shen, Z. F., & Wang, H. M. (2020). Psychological Protection Handbook for Primary and Secondary School Students and Parents During the Anti-epidemic Period. Shanghai, CHN: East China Normal University Press.

Showers, C. (1992). Compartmentalization of positive and negative self-knowledge: keeping bad apples out of the bunch. J Pers Soc Psychol, 62(6), 1036–1049. doi:10.1037//0022-3514.62.6.1036

Singh, R., & Subedi, M. (2020). COVID-19 and stigma: Social discrimination towards frontline healthcare providers and COVID-19 recovered patients in Nepal. Asian J Psychiatr, 53, 102222. doi:10.1016/j.ajp.2020.102222

Song, J. H., Colasante, T., & Malti, T. (2018). Helping yourself helps others: Linking children’s emotion regulation to prosocial behavior through sympathy and trust. Emotion, 18(4), 518–527. doi:10.1037/emo0000332

Sun, Y., & Yan, T. (2019). The Use of Public Health Indicators to Assess Individual Happiness in Post-Disaster Recovery. Int J Environ Res Public Health, 16(21). doi:10.3390/ijerph16214101

The World Bank (2021). Global Economic Prospects. https://www.worldbank.org/en/publication/global-economic-prospects/9781464816123.pdf, Accessed January.

Tibshirani, R. (2011). Regression shrinkage and selection via the lasso: a retrospective. Journal of the Royal Statistical Society Series B-Statistical Methodology, 73, 273–282. doi:DOI 10.1111/j.1467-9868.2011.00771.x

Trougakos, J. P., Chawla, N., & McCarthy, J. M. (2020). Working in a pandemic: Exploring the impact of COVID-19 health anxiety on work, family, and health outcomes. J Appl Psychol, 105(11), 1234–1245. doi:10.1037/apl0000739

Venkatesh, V. (2020). Impacts of COVID-19: A research agenda to support people in their fight. Int J Inf Manage, 55, 102197. doi:10.1016/j.ijinfomgt.2020.102197

Walsh, S. M., Chen, S., Hacker, M., & Broschard, D. (2008). A creative-bonding intervention and a friendly visit approach to promote nursing students’ self-transcendence and positive attitudes toward elders: a pilot study. Nurse Educ Today, 28(3), 363–370. doi:10.1016/j.nedt.2007.06.011

Waters, S. F., Karnilowicz, H. R., West, T. V., & Mendes, W. B. (2020). Keep it to yourself? Parent emotion suppression influences physiological linkage and interaction behavior. J Fam Psychol, 34(7), 784–793. doi:10.1037/fam0000664

Waugh, C. E. (2020). The roles of positive emotion in the regulation of emotional responses to negative events. Emotion, 20(1), 54–58. doi:10.1037/emo0000625

WHO. (2021). Coronavirus (COVID-19) Dashboard. https://covid19.who.int, Accessed 2 June.

Williams, S. N., Armitage, C. J., Tampe, T., & Dienes, K. (2020). Public perceptions and experiences of social distancing and social isolation during the COVID-19 pandemic: a UK-based focus group study. BMJ Open, 10(7), e039334. doi:10.1136/bmjopen-2020-039334

Wilson, J. M., Lee, J., Fitzgerald, H. N., Oosterhoff, B., Sevi, B., & Shook, N. J. (2020). Job Insecurity and Financial Concern During the COVID-19 Pandemic Are Associated With Worse Mental Health. J Occup Environ Med, 62(9), 686–691. doi:10.1097/JOM.0000000000001962

Wu, W. L., & Zhang, J.. (2020). Handbook of Public Psychological Protection for COVID-19. Sichuan: Sichuan Science and Technology Press.

Ye, J., Cai, S., Cheung, W. M., & Tsang, H. W. H. (2019). An East Meets West Approach to the Understanding of Emotion Dysregulation in Depression: From Perspective to Scientific Evidence. Front Psychol, 10, 574. doi:10.3389/fpsyg.2019.00574

Zhan, J., Ren, J., Sun, P., Fan, J., Liu, C., & Luo, J. (2018). The Neural Basis of Fear Promotes Anger and Sadness Counteracts Anger. Neural Plast, 2018, 3479059. doi:10.1155/2018/3479059

Zhang, S. X., Huang, H., & Wei, F. (2020). Geographical distance to the epicenter of Covid-19 predicts the burnout of the working population: Ripple effect or typhoon eye effect? Psychiatry Res, 288, 112998. doi:10.1016/j.psychres.2020.112998

Zhan, J., Ren, J., Sun, P., Fan, J., Liu, C., & Luo, J. (2018). The Neural Basis of Fear Promotes Anger and Sadness Counteracts Anger. Neural Plast, 2018, 3479059. doi: 10.1155/2018/3479059

Zhao, X. D., & Liu, Z. M. (2020). Anti-epidemic and Peace of Mind--A Comprehensive Reader for Psychological Self-help Rescue of the Epidemic. Shanghai, CHN: Shanghai Science and Technology Press.

